# Forecasting Confirmed Cases and Mortalities of COVID-19 in the US

**DOI:** 10.1101/2020.10.30.20223412

**Authors:** Babak Jamshidi, Mohsen Kakavandi, Shahriar Jamshidi Zargaran, Amir Talaei-Khoei

**Affiliations:** Postdoctoral Researcher in Biostatistics, Social Development and Health Promotion Research Center, Kermanshah University of Medical Sciences, Kermanshah, Iran; Master of Science in Mechanic Engineering, Poznan University of Technology; Master of Science in Medical engineering, Tehran University of Medical Sciences, Tehran, Iran; Associate Professor of Information Systems, Department of Accounting and Information Systems, University of Nevada, Reno, Nevada, USA

**Keywords:** COVID-19, Relative Increment, Forecast

## Abstract

**Background:** The wide spread of COVID-19 in the US has placed the country as the most infected population worldwide. This paper aims to forecast the number of confirmed cases and mortalities from 12 April to 21 May, 2020. There has been a large body of literature in forecasting epidemic outbreaks such as C algorithms with shortfall of predicting for long periods and autoregressive integrated moving average models with the limited flexibility. However, the US COVID-19 data shows great variety in the relative increments of confirmed cases. This requires a reproductive time series.

**Method:** This paper suggests a time series based on the relative increments of confirmed cases. The proposed time series assumes the changes in the time series and provides flexibility. The suggested model was applied on the data observed from 27 February to 11 April 2020 and its objective is forecasting 40 days from 12 April to 21 May 2020.

**Results:** It is expected that by May 21, 2020, the accumulative number of confirmed cases of COVID-19 in the US rises to 1,464,729, with 80% confidence interval. Our analysis also shows that by the 21^st^ of May, the cumulative number of mortalities caused by COVID-19 in the US from 18747 cases on 11 April increases to around 73250 cases on 21 May, 2020.

**Conclusion:** Our results highlight the value of reproductive strategies in time series modelling of COVID-19. Our model benefits from a reproductive strategy from a time point in which the US COVID-19 data demonstrates a sudden fall.

## 1. Introduction

An epidemic disease of unknown cause, now called COVID-19, found in Wuhan, China was first reported to the World Health Organization Office in China on 31 December 2019. Shortly after, it was found that the ongoing pandemic is an infectious disease caused by the coronaviruses, a large family of viruses that cause illness ranging from the common cold to more severe diseases such as MERS and SARS. The outbreak was declared as a Public Health Emergency worldwide on 30 January 2020. Based on its geographic spread, severity of illnesses it causes, and also its impact on society, on March 11, 2020, COVID-19 was declared as a pandemic outbreak (WHO, 2020a). As of the 8^th^ of April, 2020, the disease has spread to over 210 countries and infected 1,353,361 cases when causes the total Mortalities of 79,235 worldwide (WHO, 2020b).

The first confirmed case in the US was reported on 21 January, 2020 and on the 22nd February the first death was reported (WHO, 2020c). On the17th of March, the US reported the arrival of the disease to all the states (CDC, 2020). In one month, this number is multiplied by over 10000. As of 11 April 2020, about one-third of all confirmed cases infected by COVID-19 in the US belongs to New York. New York by almost 160 K confirmed cases, has reported the number of confirmed cases more than all countries (other than the US) (CDC, 2020). It is noticeable that up to now, USA has faced the greatest new mortalities (2035) and new confirmed cases (34196) in one day on 10 and 4 April, respectively. In addition, Active cases in USA on 11 April were 481849 which is approximately equal to active cases of the other eight countries with the most confirmed cases (Spain, Italy, France, Germany, China, United Kingdom, Iran, Turkey) (WHO, 2020c)

We use the US recent COVID-19 data from 27 February to 11 April 2020 to model and forecast the development of the outbreak in the US for the period of April 12th – May 21st, 2020 predicting the relative increment of the confirmed cases and Mortalities using two-part time series. The first part represents the propagation of the disease in the first period in which the rate of transition is high. The second part models an irreversible fall in the relative increment and continues the extinction of the disease.

There has been a body of research presenting forecasting trends in fields such as epidemiology (Chretien et al., 2014; Paul et al., 2014; Talaei-Khoei et al., 2019), economics (Baltagi and Baltagi, 2001; Scott and Varian, 2015), weather (Kumar and Jha, 2013; Radzuan et al., 2013; Voyant et al., 2013). Forecasting epidemic outbreaks plays a significant role in the success of epidemiology and public health systems to effectively respond to the epidemic situations. As such, there are several attempts suggesting methods for forecasting the confirmed cases in outbreaks. Chretien et al. (2014) reviews the methods in forecasting influenzas in human populations. From theoretical standpoint, forecasting methods are provided with time series data from previous stages of an outbreak and predicts surveillance attributes of a disease such as the confirmed cases in future. In order to do so, there have been several statistical methods (Reis and Mandl, 2003; Rounds et al., 2017; Zeger et al., 2006; Zhang et al., 2014) suggested by literature. Accordingly, there have been several attempts by professional bodies. For example, The Centres for Disease Control and Prevention initiated a challenge to forecast the 2013-2014 United States influenza season (Biggerstaff et al., 2016). Similar challenges was also organized for Ebola (Viboud et al., 2018), and Dengue (NOAA, 2016). The literature in time series forecasting has used aberrancy-detection algorithms to identify temporal changes in the data. These temporal changes may be the indicator of epidemic outbreaks (Murphy and Burkom, 2008). The Early Aberration Reporting Systems from Centres for Disease Control and Prevention utilizes C algorithms. However, C algorithms suffer from short-term prediction capabilities (Tokars et al., 2015). General speaking, aberrancy-detection algorithms focuses on a single time point in a given time when the outbreaks occurs and they are limited to be used to forecast the number of confirmed cases in the future of outbreak (Kass-Hout et al., 2012). Zhang (2003) suggests the use of Autoregressive Integrated Moving Average (ARIMA) to address the shortfall of aberrancy-detection algorithms. ARIMA models have been largely used in forecasting infectious diseases such as dengue (Wongkoon et al., 2012) and tuberculosis (Rios et al., 2000). ARIMA models are committed to the assumption that in the Autoregressive model under the study, the present value of the time series is a linear function of the past values and random noise (Akaike, 1969). Second, the present value of the time series is a linear function of present and past residuals in the moving average model (Haining, 1978). Third, ARIMA is based on both Autoregressive and moving average as well as the past values and residuals (Rojas et al., 2008). These assumptions limit the flexibility of ARIMA models to reproduce the different stages of time series.

> **Looking at data from COVID-19, it appears that the outbreak has demonstrated decreasing relative increment in the number of new cases. Since ARIMA models focuses on the linear relationships between the present values with past data and results, the forecasting ability of ARIMA models for COVID-19 would be limited. Addressing the shortfall of ARIMA models, this paper, using a novel model to reproduce the time series of COVID-19, aims to forecast the number of new cases in the US. The present paper adopts a stochastic approach due to two main reasons. Firstly, there are several sources of uncertainty in a pandemic outbreak (Chowell et al., 2020) such as COVID-19. Randomness is a major player particularly at the beginning of an outbreak. Secondly, the stochastic approach used in our model promotes the flexibility of the model. The proposed method commits us to three assumptions. Firstly, the relative increase in the number of confirmed cases are increasing at the beginning, then it experiences a sudden fall and it decreases thereafter. Looking at the data from COVID-19 in the US, we can confirm that this is the case, See** Day 0: 27 February 2020, Day 45: 12 April 2020.

Figure 1. Secondly, at any time, the time series of relative increase in the confirmed cases holds a normal distribution. The COVID-19 in the US demonstrates a normal distribution for the relative increase of the confirmed cases. Third, over time, the ratio of variance to mean remains constant, which is the case for COVID-19 in the US.

**Figure 1.**
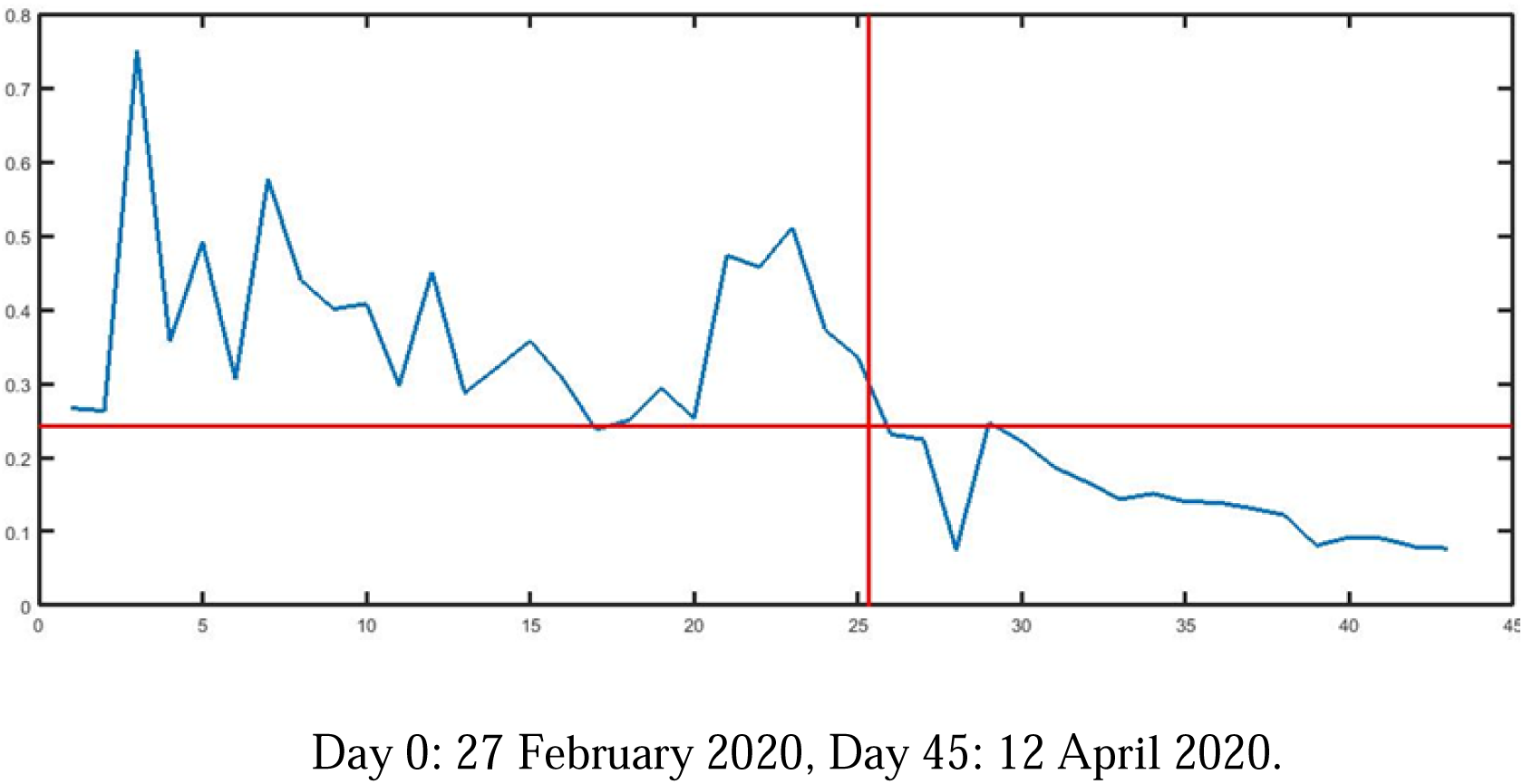
The time series of relative increments and the time of passing from the first stationary period to the new decreasing period.

The present paper relates to the recent articles attempting to model the spread of the COVID-19 19 (Berger et al., 2020; Kucharski et al., 2020; Read et al., 2020). The present study differs from these papers from Data and Method perspectives. Berger et al. (2020) study the data from the United States and it investigates the role of testing and quarantine as two effective practices to contain the disease. The article does not use time series analysis. Kucharski et al. (2020) look into cases in Wuhan and internationally exported cases from Wuhan. The article addresses uncertainty of COVID-19 by using some outside databases, while in this study we only utilize data from the US. Read et al. (2020) explores the early data from Wuhan and focuses on epidemiological parameter estimates. However, the present paper mainly looks at time series analysis to forecast the newly confirmed cases.

The rest of this paper is organized in the following way, Section 2 presents the proposed method. Section 3 presents the results of our forecast. Section 0 discusses the paper and provides some limitations as well as directions for future research.

## 2. Methods

In this section, we will discuss the proposed model, and the estimations. We will also simulate the model and the estimation on the data from the US to forecast for the next 40 days. The calculations, simulations have been conducted in MatLab R2015b. In addition, we have opted five output forecasting variables to compare, namely the average curve and 80% upper bound, 80% lower lower bound and two realizations. All these outputs are computed based on 100 times simulations of the fitted models.

The data in this section is obtained from WHO situations reports (WHO, 2020c) where we collected the data for the US from 27 February to 11 April 2020 and the paper forecasts from 12 April to 21May, 2020.

### 2.1 Definition of Relative Increment of Confirmed Cases and Mortality Rate

#### Relative Increment of Confirmed Cases

Let *Y*_*t*_ is a time series of the number of confirmed cases up to time *t*. We aim at studying 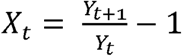, called the daily relative increment of confirmed cases.

#### Mortality Rate

In order to define the mortality rate of COVID-19 in the US, we define 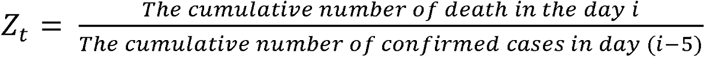.

### 2.2 Proposed Time Series

The model we applied has five positive parameters (*b,lR,K,θ,a*); For, *t* = 1,…,*b* 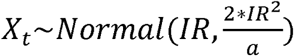 which confirms the second assumption of our time series, See Section 1.

1. For = *b* + 1, *b* + 2, … 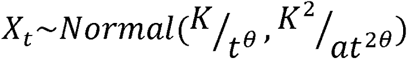, where;
  - *b*: The length of the first periods of the spread of the disease,
2. *IR*: The geometric mean of the relative increment in the first periods of the spread of the disease,
3. *θ*: The acceleration of falling of the relative increments after the first days of spreading (*X*_*t*_ ∝ *t*^−*θ*^ *for t* = *b* + 1,*b* + 2, …),
4. *a* : The fixed ratio of the mean to the variance (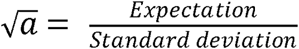), which confirms the third assumption of our time series, See Section 1.
5. *K*: The adjusting coefficient for the curve *t*^−*θ*^ to fit the time series of the relative increment after the first period of the spread.

The estimation of the proposed time series for the US Data of COVID-19 would be discussed in the next subsection.

### 2.3. Estimation for Increment of Confirmed Cases of COVID-19 in the US

In this section, we discuss on how the time series explained above will fit to the COVID-19 data from the US. To estimate the parameters of the model, the following practices must be taken:

- take *b* as the first point that the geometric mean of the relative increments in the previous points exceeds 3/2 times the geometric mean of the next three points.

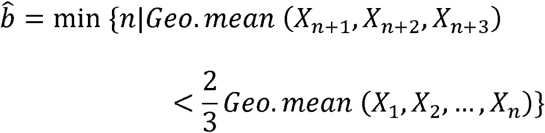

which can be computed as

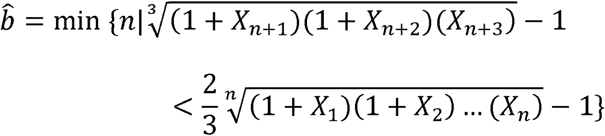

Graphically, this time can be identified as the time when the plot of relative increments falls irreversibly, see Day 0: 27 February 2020, Day 45: 12 April 2020.

Figure 1.

Day 0: 27 February 2020, Day 45: 12 April 2020.

Figure 1 presents the relative increment in time series of the COVID-19 confirmed cases for the US. The figure demonstrates that the time series has a time point in which the first stationary period ends, and the second decreasing period starts. In this case, it is resulted that 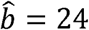. This confirms our first assumption for the proposed time series, See Section 1.

- calculate the geometric mean of the ratio cumulative numbers in the previous points (1+*X*_*t*_) from, *t* = 1 to *t* = *b* as the estimation of the parameter *IR* as 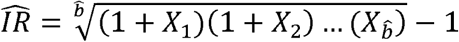. For the US COVID-19 data, 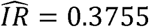.
- estimate the parameters *θ* and *K* due to the following linear relation

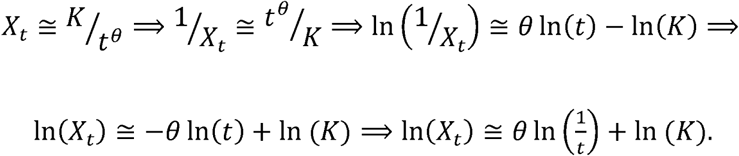

For the US COVID-19 data, the calculations lead in 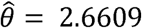 and 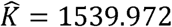, See Figure 2.
- Multiply all the observations after, *t* = *b* by 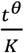 to have an identical mean and variance for all the newly obtained data (*W*_*t*_)

**Figure 2.**
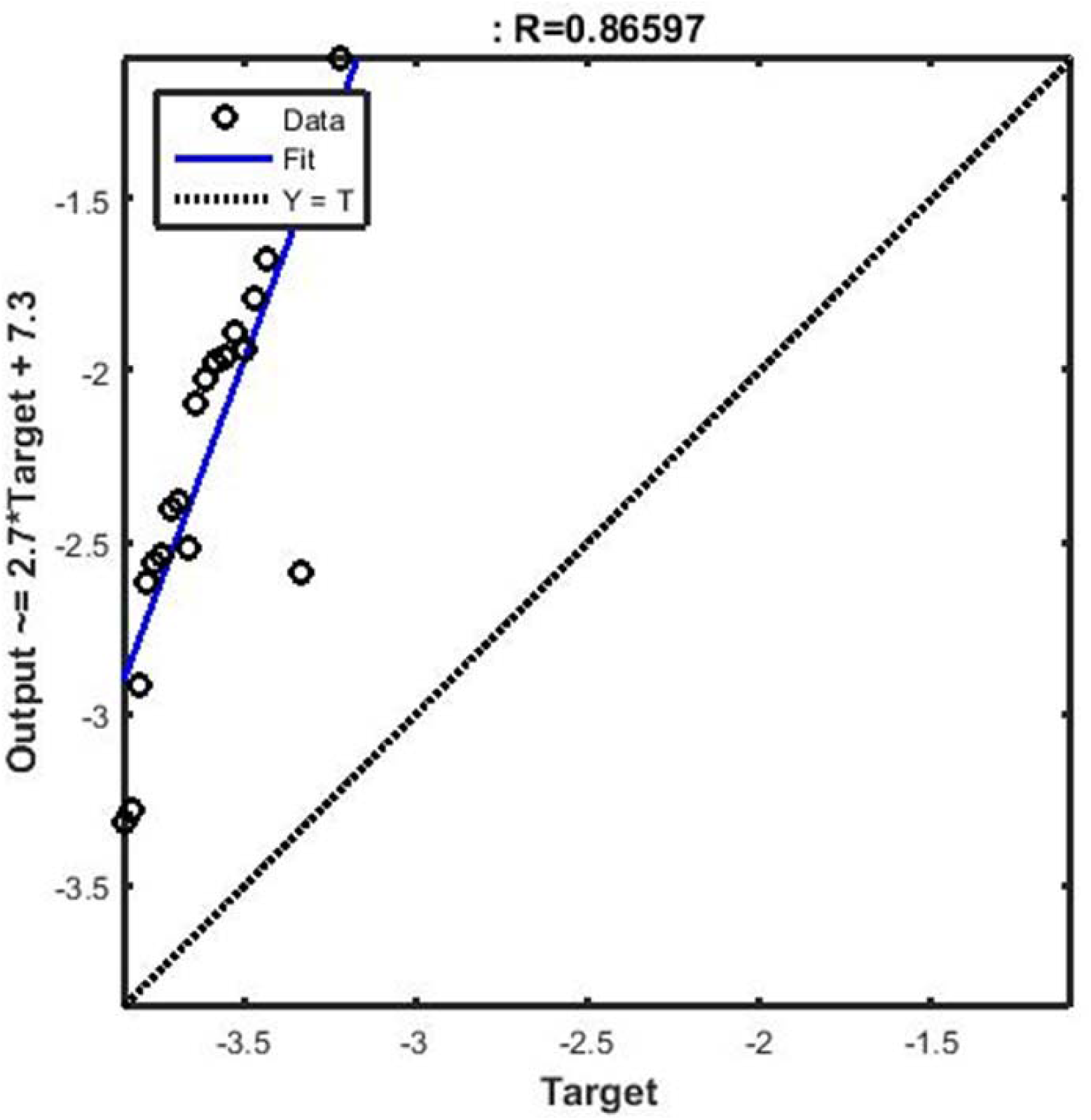
Estimating the linear regression for fitting the association between relative increment and the inverse of time to obtain estimations of the parameters θ and K.

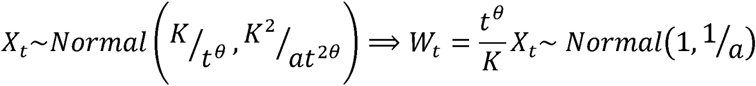

Therefore, the variance of the newly recorded data is a good candidate for estimating 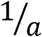. Accordingly, 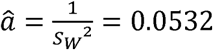.

> **All in all, the US COVID-19 data from 27 February to 11 April 2020 generates the estimation of** 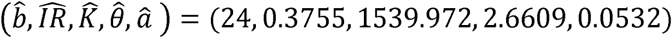 **to simulate and predict the US confirmed cases using the time series introduced in Section 2.2**. Day 0: 22 March 2020, Day 20: 12 April 2020.

Figure 3 demonstrates that our model has an appropriate fitting power to the relative increments of the of the confirmed COVID-19 cases in the US. Based on the model, the relative increment is decreasing from around 7% to about 1% (its 80% confidence interval is 0.88% - 1.56%).

**Figure 3.**
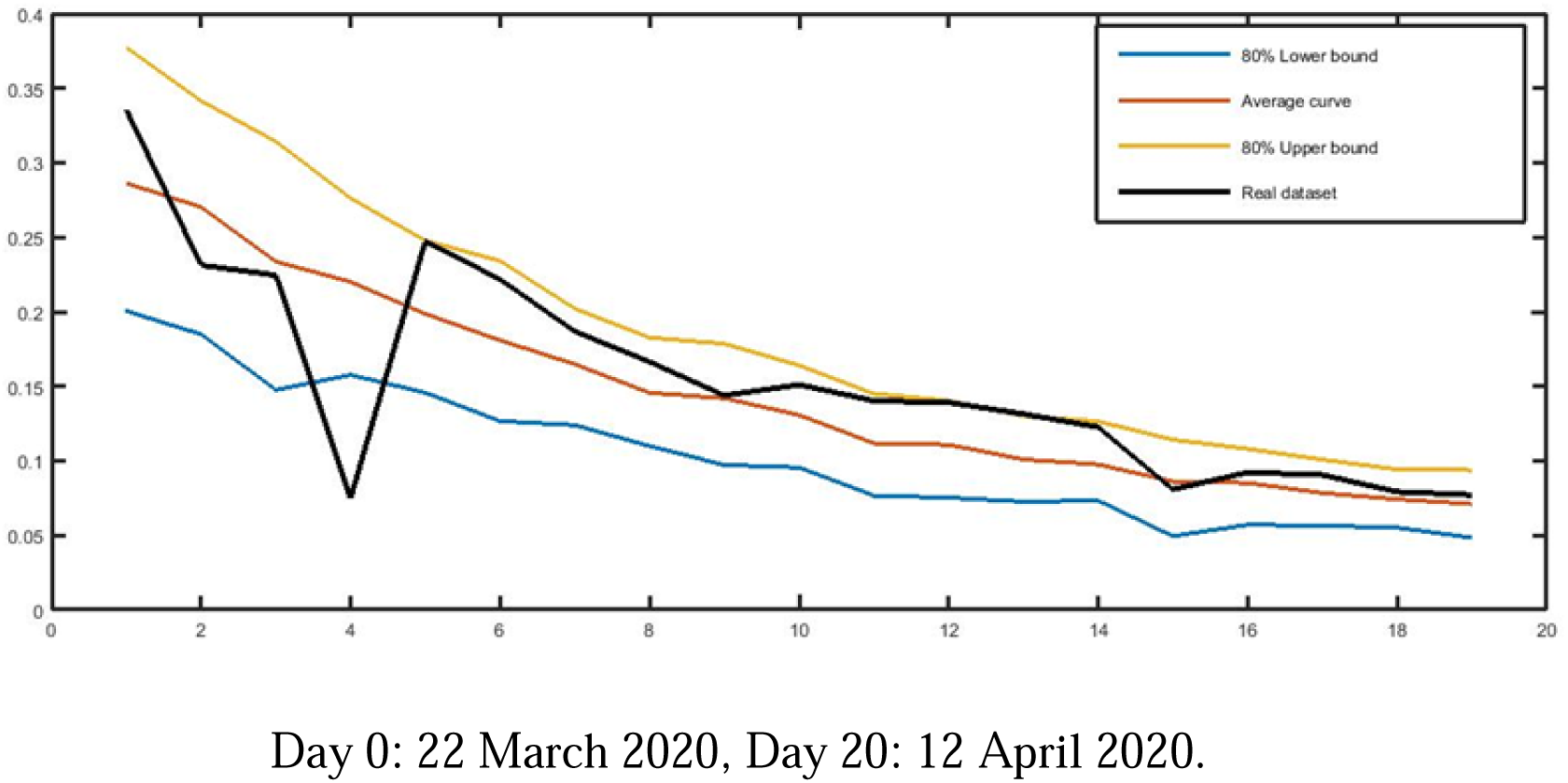
Fitting Power of the model to the relative increments of confirmed COVID-19 cases in the US.

**Figure 4.**
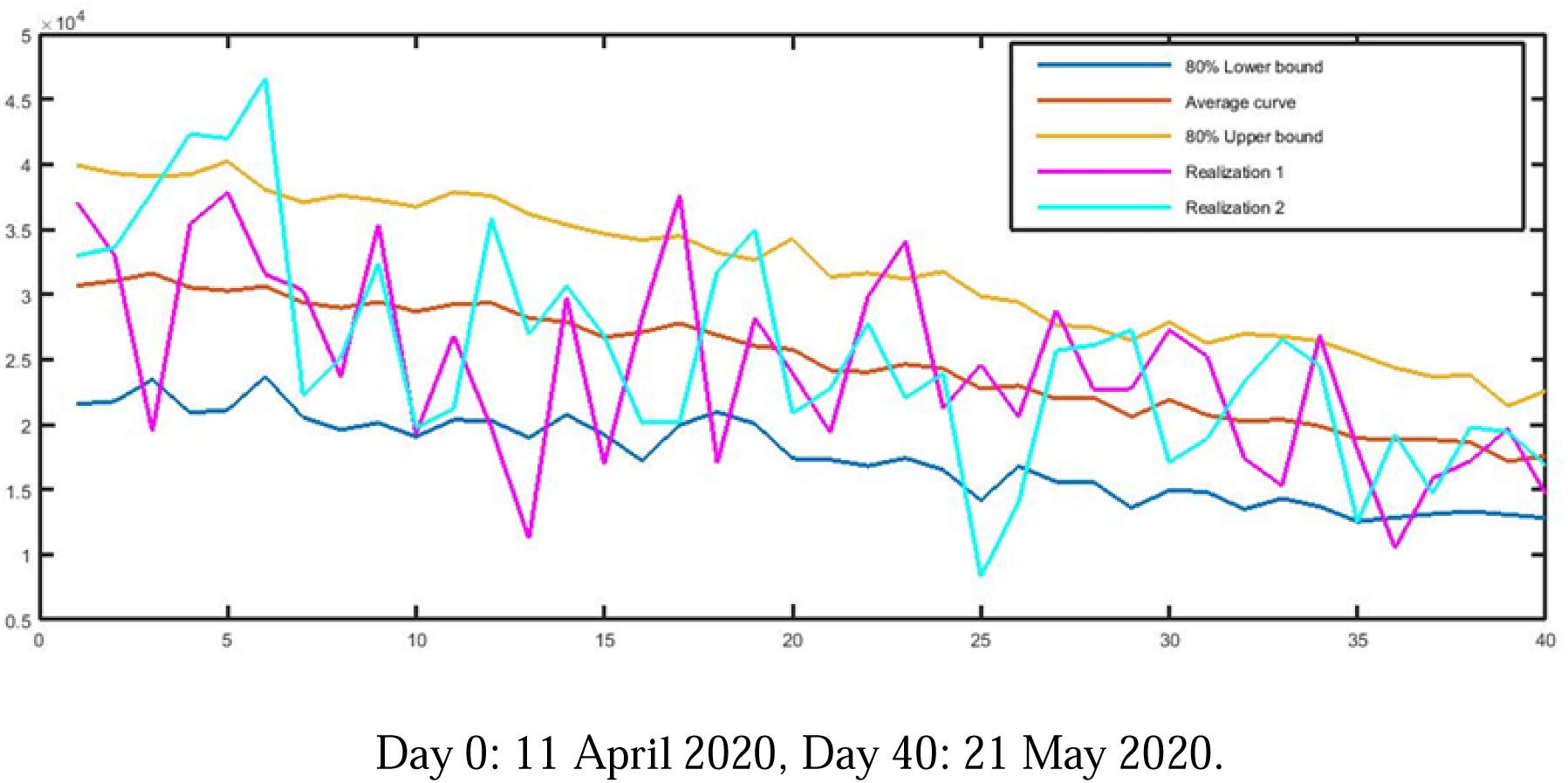
Forecast of the relative increments of the COVID-19 cases in the US, from 12 April to 21 May, 2020.

**Figure 5.**
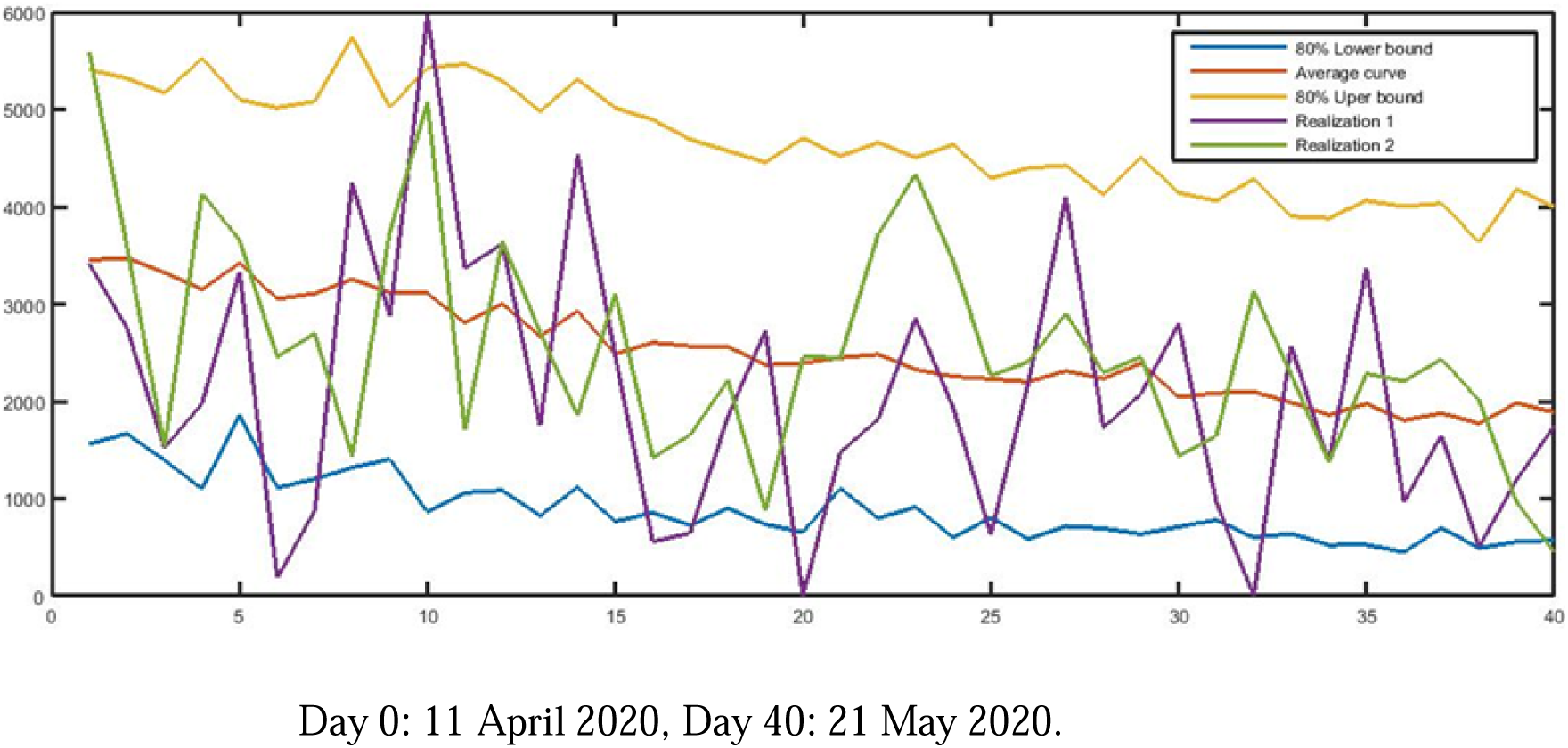
Forecast of the newly confirmed cases of COVID-19 in the US, from 12 April to 21 May, 2020.

**Figure 6.**
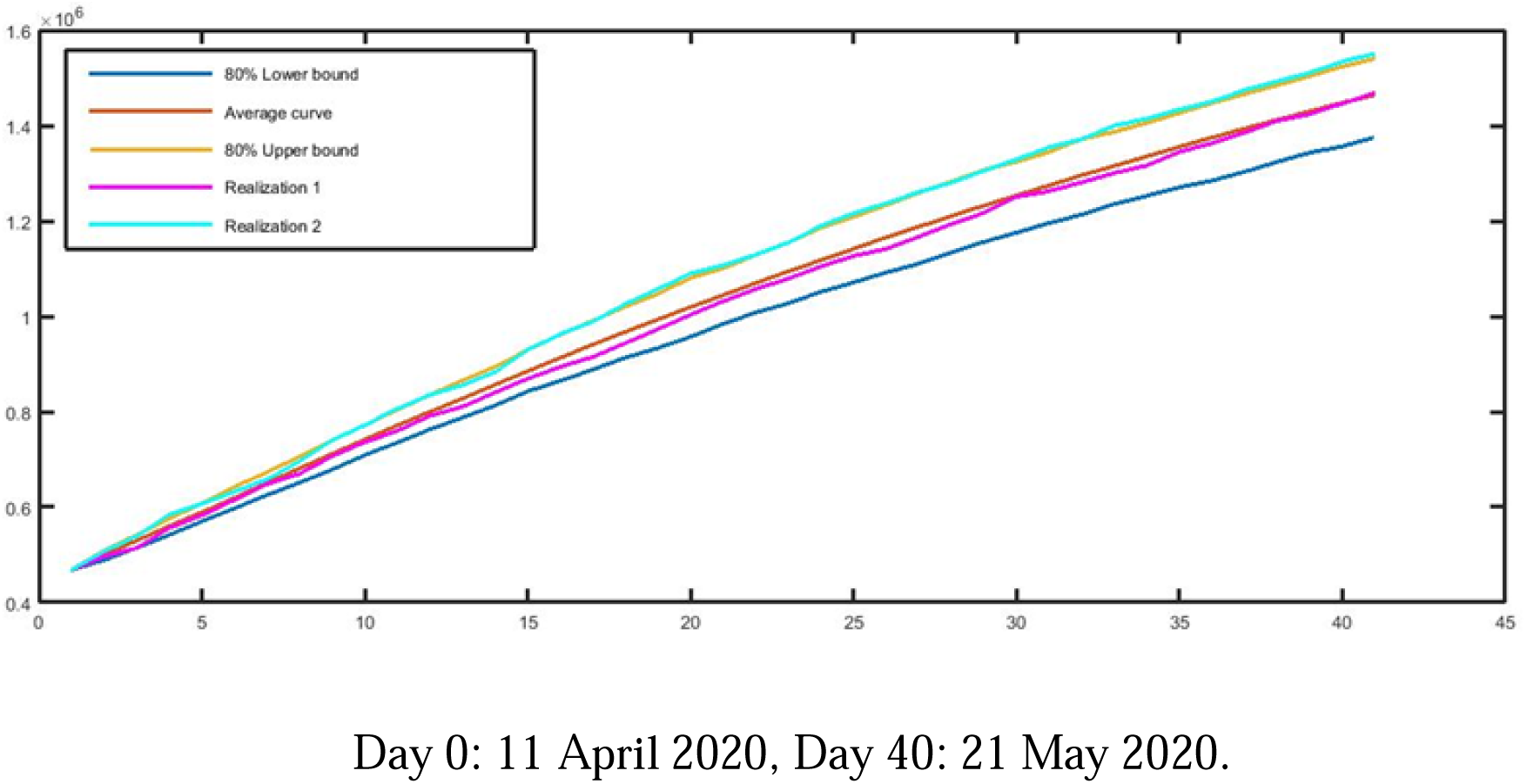
Forecast of the cumulative number of confirmed cases of COVID-19 in the US, from 12 April to 21 May, 2020.

**Figure 7.**
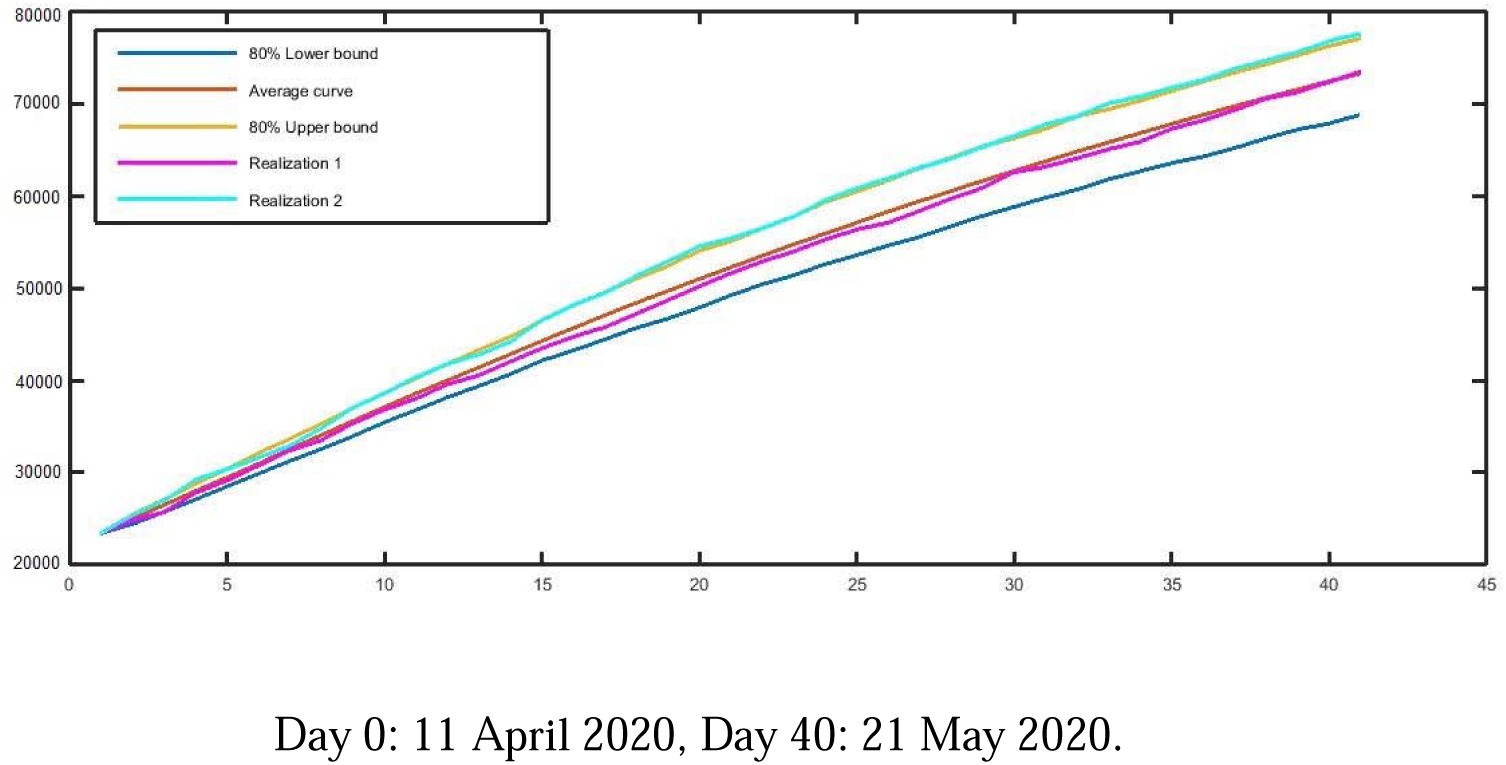
Forecast of the cumulative number of mortalities caused by COVID-19 in the US, from 12 April to 21 May, 2020.

### 2.4 Estimation for Mortality Rate of COVID-19 in the US

We utilize the morality rate of 5% to estimate the number of deaths in the US from 28 February to 11 April.

## 3. Forecasting Results

In this section, we first forecast the relative increments of confirmed COVID-19 cases in the US from 12 April to 21 May, 2020. Accordingly, we will forecast the accumulative number of conformed cases in the next 40 days in the US. Second, we forecast the COVID-19 mortality rate and accordingly the accumulative number of Mortalities from 12 April to 21 May, 2020. In our forecast, as mentioned earlier, we use five different forecasting output variables, namely 80% lower bound, 80% upper bound, average and two possible realizations.

### 3.1 Forecasting relative increment and accumulative number of confirmed cases

Day 0: 11 April 2020, Day 40: 21 May 2020.

Figure presents our five forecasting variables for the relative increment of confirmed cases of COVID-19 in the US from 12 April to 21 May, 2020, presented on the model as day 0 to day 40.

Figure presents the forecast of the newly confirmed cases of COVID-19 in the US from 12 April to 21 May, 2020. As demonstrated by Figure, based on the decreasing trend of daily relative increments, despite the increasing cumulative number of confirmed cases, the new confirmed cases infected by the pandemic follows a decreasing pattern. In the first 10 days of April, the variable is fluctuate around 30000 while our prediction says that this variable decreases to [12801 22578] with the probability of 80% (the point prediction is equal to 17551) on 21 May 2020.

Figure presents the forecast of cumulative confirmed cases for the studied period. The forecast predicts that over the period of 12 April to 21 May, 2020, the cumulative number of cases rises from 502876 on 11 April to 1464729 cases on 21 May 2020, with 80% confidence interval equal to [1375362 1540424].

### 3.1 Forecasting accumulative number mortalities

According to Figure, the number of Mortalities caused by COVID-19 in the US increases from18747 cases on 11 April to around 73000 cases on 21 May, with 80% confidence interval equal to [69 77] K. It means we will encounter with about 1350 daily Mortalities on average over the studied period. The slope of the increasing graph for the cumulative number of Mortalities is falling as well as the graph of the cumulative number of confirmed cases.

## 4. Conclusions, Discussion and Limitations

The relative increments in confirmed cases of COVID-19 in the US has shown in the beginning an increase in the rate of its growth, then it has experienced a sudden fall and continued to decrease in a relaxer rate. This demonstrates a great variety and presents different stages of COVID-19 in the US. This paper aims to forecast the number of confirmed as well as the Mortalities of COVID-19 in the US from 12 April to 21 May, 2020. Given the shortfall of C algorithms to forecast in longer terms and limited flexibility of ARIMA to model different stages of time series in an outbreak like COVID-19, we suggested the use of relative increment measure for the confirmed cases and the mortality rate for the number of Mortalities caused by COVID-19. The model presented in Section 2 is able to model COVID-19 data in the US because the proposed structure takes variety of different stages in the data.

Applying the model to the US COVID-19 data from (WHO, 2020c), the paper predicts 1464729 confirmed cases on the 21^st^ of May, 2020. The forecast illustrates that the US can be considered the first country to face the problem of the explosion of the pandemic for COVID-19. Although the accumulative number of confirmed cases in the US will be increasing to the 21st of May, the forecasting exercise presented in this paper found some evidence that the relative increment of the cases is falling. This shows the growth rate of confirmed cases of COVID-19 in the US will be dropping. The estimated drop in the relative increments is driven by the changes in behaviours of population or specific social distancing measures that the US has implemented.

> **Our results highlight the value of reproductive strategies in time series modelling of COVID-19. For instance, the rapid growth of COVID-19 confirmed cases in the US at the beginning of the outbreak would result into inaccurate forecasts, while** Day 0: 27 February 2020, Day 45: 12 April 2020.

Figure 1 demonstrates a sudden fall followed by a relaxer decreasing rate of growth. This problem has been also reported by Kucharski et al. (2020). Our model benefits from a reproductive strategy from a time point in which the US COVID-19 data demonstrates a sudden fall.

The model presented in this paper is limited to three assumptions on the relative increments. Firstly, similar to Italian COVID-19 it must rapidly grow with a dramatic increasing rate, experiences a sudden fall and then decreases in a smaller rate. Second, it must follow a normal distribution and third, over time, the ratio of variance to mean remains constant. Although holding these assumptions is an advantage of the proposed method to model the US COVID-19, it may limit the generalizability of the model fitting to other outbreaks.

Researchers in this area are recommended to apply the method to COVID-19 data from other regions. Our initial analysis shows the model fits to Iranian. UK and Italian data. From operational perspective, in order to simplify the proposed model for quicker implementation of forecasts, the model can be simplified by removing the two first parameters suitable to describe the gradual decrease of the relative increments (*t*=*b,b*+1,…).

## Data Availability

N/A

https://www.worldometers.info/coronavirus/coronavirus-cases/

## Summary Points

1. The study aims to forecast the number of confirmed cases of COVID-19 in the US.
2. The paper builds a time series model based on COVID-19 data from 27 Feb to 11 April 2020.
3. The study forecasts 1,464,729 confirmed cases of COVID-19 by the 21st of May, 2020.
4. The paper forecasts the accumulative number of mortalities of COVID-19 to around 47000 on the 21st of May, 2020.
5. The proposed method is based on relative increments of confirmed cases.

